# Task-related Aperiodic EEG (1/f) Activity in Autism

**DOI:** 10.1101/2025.10.16.25338172

**Authors:** Magdalena Matyjek, Salvador Soto Faraco, Claudia Alvarez Martin, Mireia Torralba Cuello

## Abstract

Autism has been hypothesised to involve atypicalities in the balance between neural excitation and inhibition (E/I). Aperiodic EEG activity, characterised by the 1/f exponent of the power spectrum, provides a proxy of cortical E/I dynamics, yet prior studies in autism report mixed findings. One potentially important modulator of EEG slope is task engagement. Here, we examined variations in aperiodic slopes in autistic (n = 35) and neurotypical (n = 39) adults during passive viewing and an active, goal-directed task. The data revealed that autistic participants exhibited steeper slopes (indicative of increased inhibition vs. excitation) during the active task relative to passive viewing, whereas neurotypical participants showed no significant task-related changes. These findings suggest that aperiodic activity reflects dynamic, task-dependent neural adaptation rather than baseline group differences. Task engagement may reveal compensatory inhibitory processes in autistic adults, underscoring the importance of considering task demands and individual variability when investigating E/I balance in autism.

## 1. Introduction

For two decades, it has been hypothesised that autism spectrum disorder (autism) involves to atypicalities in the balance between neural excitation and inhibition (E/I) ^1,2^. Excitation, predominantly driven by glutamatergic input, increases neurons likelihood to fire and respond to stimuli, whereas inhibition, mediated mainly by gamma-aminobutyric acid (GABA), prevents runaway activity leading to epileptogenesis ^3^. The original model of E/I imbalance in autism hypothesised elevated cortical excitation relative to inhibition, potentially explaining sensory hyper-responsivity and other autistic characteristics ^2^. Early support came from observations of reduced GABAergic activity and the frequent co-occurrence of epilepsy in autism ^2,4^. Since then, a large body of work further refined this hypothesis. Animal models showed consistent evidence that E/I imbalance affects autism-relevant behaviours ^5^, whereas human evidence demonstrated that this imbalance varies with age, brain region, and method used to measure E/I ^3,4,6,7^. Overall, findings in humans remain mixed, with some evidence pointing to overexcitation and some to overinhibition in autism ^4^.

A powerful approach to study the E/I neural systems is electroencephalography (EEG), which allows the estimation of both the periodic (ongoing oscillatory rhythms and time-locked evoked responses) and the aperiodic (scale-free) neural activity ^8,9^. In the power spectrum, oscillations correspond narrowband peaks, whereas aperiodic activity is a broadband signal whose amplitude typically decays with frequency, following a power law that has been related with cortical excitability and E/I balance ^10,11^. For example, computational and animal studies show that increasing net inhibition (lower E/I) steepens the slope of aperiodic activity, whereas increasing cortical excitation (higher E/I) flattens the slope ^11^. Although aperiodic activity used to be treated as “noise”, at present there is ample evidence of its relevance for human cognition. For example, steeper slopes of aperiodic activity have been related to increased vigilance states in awake population ^12^, faster cognitive speed ^9,13^, and stimulus processing compared to baseline period ^14^. Importantly, alterations in the slope of aperiodic activity have been associated with some neurological disorders: compared to control populations, steeper slopes have been found in Parkinson Disease, disorder of consciousness, and in epileptic patients previous to seizure onset, whereas flatter slopes have been observed in attention deficit hyperactivity disorder ^1^. In autism, evidence regarding alteration in aperiodic activity slopes is mixed ^1^.

In children, effects in both directions (steeper or flatter slopes) have been observed in autistic compared to non-autistic individuals. On the one hand, Carter Leno and colleagues ^15^ found *steeper* slopes in aperiodic activity —indicating increased inhibition with respect to excitation— at 10 months correlated with higher levels of autistic traits at 36 months, but only in children with lower executive attention ability. This suggests that higher executive functioning skills may absorb the negative impact of early cortical atypicalities. Similarly, preterm infants who later showed higher scores on early autism screening measures tended to have *steeper* EEG slopes in the 1 to 20 Hz range at birth ^16^. On the other hand, Manyukhina et al. ^17^ reported that autistic boys with below-average IQ showed *flatter* aperiodic slopes (suggesting hyper-excitation) than either typically developing boys or autistic peers with typical IQ.

It is also unclear whether slope atypicalities in autistic individuals persist into adulthood. Li and colleagues ^18^ measured resting-state EEG with eyes closed in adults and found the aperiodic slope to be significantly flatter in the autism group, but this effect disappeared once age was controlled. Dede et al. ^19^ systematically tested hundreds of features in large resting EEG datasets of autistic and neurotypical participants (n=776). They found almost no replicable differences between the groups, including the aperiodic slopes (although age and sex predicted 40% and 16% of EEG variance respectively). Thus, so far published resting-EEG studies of autistic adults report either null effects or age-confounded trends in the slope of aperiodic activity.

Although these mixed findings might cast some doubt on the relationship of E/I balance with autism, and/or the sensitivity of EEG to capture relevant E/I patterns, some considerations are in point. The sensitivity of the aperiodic slopes as a proxy might be moderated by some participant characteristics and task context. For instance, the steepness of the aperiodic slope in EEG is known to negatively correlate with age ^20,21^. In addition, heightened cognitive demand (i.e., when performing a demanding task) is known to elevate neural excitability and therefore render flatter aperiodic slopes ^10^. A case in point is that whereas EEG slope is steeper during eyes-closed rest than in eyes-open conditions both in autistic and typically developing children ^17^, autism-related differences tend to emerge when engaged in active tasks. Hill et al. ^22^ report that during emotional face processing, higher autistic traits were associated with steeper exponents, suggesting a more inhibitory-like spectral profile under increased cognitive demand. Some neuropharmacological studies further highlight that group differences are revealed in context-dependent changes, rather than simply as baseline differences. Ellis et al. ^23^ reported that when autistic and non-autistic adults received low or high doses of a GABA agonist (resulting in increased central inhibition), only the autism group showed steeper slopes under low-dose administration, whereas neurotypical participants displayed either no change (eyes closed) or flatter slopes (eyes open), relative to baseline (placebo). One interpretation is that steeper slopes in autism may reflect homeostatic compensation for heightened excitation in order to stabilise E/I balance ^15^. Taken together, studies including active tasks suggest that aperiodic slope is not a static marker, but rather a dynamic measure; autism-related differences emerge in how activity adapts between passive and active states. An open question, therefore, is whether autistic and neurotypical groups diverge specifically in the adjustment of aperiodic activity across different task demands.

To answer this question, in the present study we aimed to test for variations in aperiodic slope between autistic and non-autistic adults considering neural activity as participants engaged in passive viewing vs. active goal-directed task conditions. To this end, we analysed the aperiodic slope of the EEG signal in data from a previous (unrelated) study of our group ^24^. In the active task, participants were asked to identify spoken words produced by actors from video recordings. In the passive condition, participants watched static pictures of the same actors but had no task to perform. According to prior findings ^14^, we expected to observe an overall effect of task with steeper slopes (higher net inhibition) in active vs passive viewing. Further, we predicted a significant task by group interaction, which would indicate group differences in dynamic E/I adaptation to task demands. However, we did not have predictions for the direction of the interaction given the inconclusive results in the literature so far. In addition, given that aperiodic slopes flatten with age ^10,21^ and both age and sex are important predictors of autistic vs. neurotypical group differences in EEG signals ^19^, we planned to explore the data with and without controlling for these variables in the analysis.

## 2. Methods

### 2.1. Participants

Sample characteristics are summarised in Table 1. The study was approved by the Institutional Committee for Ethical Review of Projects at University Pompeu Fabra (CIREP-UPF; No. 258). All participants were Spanish-speaking volunteers between 18 and 60 years of age, with no intellectual disability and with normal or corrected-to-normal vision and audition. All autistic participants provided a written diagnosis of Autism Spectrum Disorder previously made by a professional. Non-autistic participants were excluded if they had any history of neuro/psychological/psychiatric disorders. All read study information and gave a written consent to participate in the study. Details on recruitment can be found in our previous publications with this cohort ^24,25^.

**Table 1.**
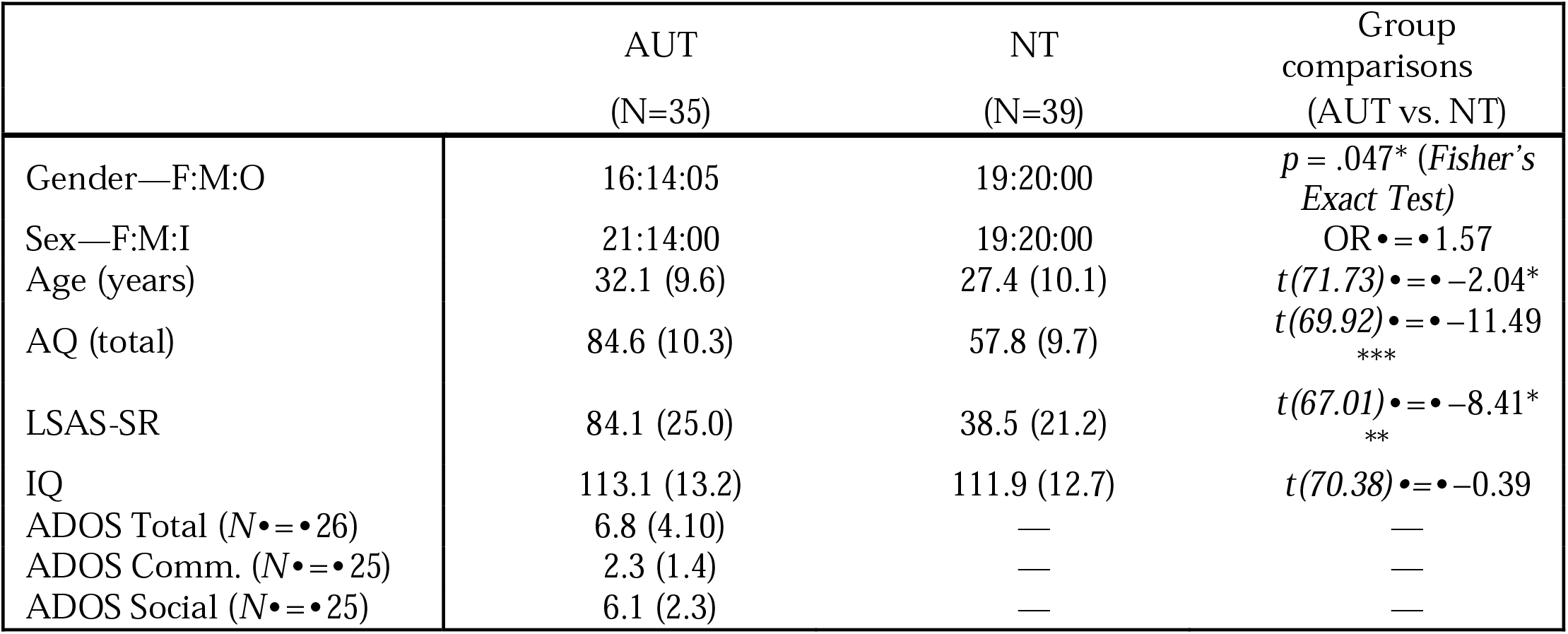
Demographic and trait characteristics of the sample. Count is provided for gender and sex, and means (with standard deviations) for all other items. F/M/O/I•=•female, male, other, intersexual; AQ•=•Autism Spectrum Quotient 10-item Spanish version ^26^>; LSAS-SR•=•Liebowitz Social Anxiety Scale-Self Reported ^>27^>; IQ•=•Intelligence Quotient measured with Raven’s Progressive Matrices 2 ^28^; ADOS=Autism Diagnosis Observation Schedule 2^29^; OR•=•odds ratio in Fisher’s Exact Test. Statistically significant tests were marked with ***for p•<•0.001.

### 2.2. Task

Participants performed two tasks: first passive viewing and then an active task (the order was always the same). The passive task consisted of watching a sequence of static pictures for 2 s of three actors randomly selected from a pool of four (15 trials per actor). In the active task, participants watched the same actors uttering verbs and were asked to identify the produced word. In this task, each of the 61 verbs was presented in three conditions: audio-visual (actors both heard and seen), visual (actor’s microphone “malfunctioning”), or auditory (camera “turned off”). See a detailed description of this task in ^25^.

We analysed aperiodic slopes in the EEG signal from all trials in the passive task (45 trials per participant) and from the visual-only condition of the active task (64-65 trials per participant). Please note the original dataset also contained trials with auditory speech (auditory only and audio-visual conditions) but these trials were not included here because they are not comparable to the passive (silent) condition. This also ensured a more balanced comparison in terms of trial numbers across tasks. Please note that the autistic and neurotypical groups did not significantly differ in terms of the accuracy of word detection or in terms of brain responses (alpha suppression) induced by the active (visual-only) condition (see details in ^24^).

### 2.3. EEG data acquisition and processing

EEG data were recorded continuously at 500□Hz using Brain Vision Recorder and 61 active electrodes (actiCHAmp, Brain Products GmbH) positioned according to the extended 10–10 international system on an elastic cap, with AFz as ground. Impedances were kept below 20□k•. Eye movements were monitored via electrodes placed at the right outer canthus (HEOG) and below the left eye (VEOG). The tip of the nose served as the online reference during acquisition.

Offline preprocessing was carried out in Matlab (Mathworks) with EEGLAB toolbox ^30^. Channels exhibiting flat signals or abnormally large voltage fluctuations (as judged by visual inspection) were discarded. Data were re-referenced to the average reference. A high pass frequency filter of 1Hz was applied only at this point of the analysis, and timepoints of up to 2ms that showed power values higher than 300 •V were discarded. Blink and eye-movement related activity was detected and corrected using independent component analysis ^31^ and package IClabel ^32^. A mean of 2.44± 0.87 components were discarded per subject. Discarded channels were interpolated using spherical splines based on neighbouring electrodes. Data were epoched in segments of variable duration, from the start to the end of the video. Only segments with a duration > 2 s were retained for further analysis. For the passive task, 35±10 epochs 2.01±0.04 s long were created (37±10 epochs 2.00±0.02 s long for NT and 34±9 epochs 2.02±0.05 s long for AUT). For the V condition in the active task, 57±13 epochs 4.7±0.8 s long were created (61±14 epochs 4.7±0.4 s long for NT and 52±17 epochs 4.7±0.8 s long for AUT).

### 2.4. Aperiodic activity analysis – FOOOF algorithm

EEG spectral analysis was performed using Fieldtrip ^33^. Fourier Transform (FT) was applied to epochs zero-padded to 20 ms (frequencies of interest 1 to 1000 Hz in steps of 0.25 Hz). Power spectrum obtained with FT was averaged across electrodes, resulting in one spectral estimation per trial. The *FOOOF* package for Matlab ^10^ was used to characterize aperiodic activity. Aperiodic slope was estimated in the 4 to 50 Hz range ^34^, no limit to the number of detected peaks was set, the minimum height of detected peaks was 0 dB. In order to discard defective estimations of the slope, a threshold was set for the goodness of fit (R) of aperiodic activity. Only estimations with R>0.8 were accepted. In addition, slope values implying larger power values at high frequencies compared to low frequencies were also discarded. For the passive task, 32±12 slopes per subject (33±10 for the NT and 32±15 for the AUT) were included in the analysis. For the active task, 50±17 slopes per subject were included in the analysis (55±14 for the NT and 44±19 for the AUT). Given that the length of passive trials was shorter than active task trials, we checked that slope estimation was not biased by segment length (see Supplementary Materials section 5).

### 2.5. Statistical analysis

All statistical analyses were performed using R v.4.4.2 ^35^. The significance level for all the tests was set to alpha=0.05, unless otherwise noted.

We analysed the aperiodic slopes using multiple regression models with mixed effects (random intercepts for subjects) with the *lmerTest* package v.3.1–3 ^36^. First, to test whether slopes were flatter in active vs passive tasks (first prediction), and second to test for an interaction between task and group (second prediction). To test these predictions, we built a basic model with (single trial) aperiodic slopes as the dependent variable, using group (autistic and neurotypical; AUT and NT), task (active and passive), and their interaction as independent variables:

#### Model 1

Slope ∼ Group * Task + (1 | ID).

In a second model, we explored the effects of biological sex (Female, Male), age, and their interactions with Group and Task:

#### Model 2

Slope ∼ Group * Task * Age * Sex + (1 | ID).

The models were first checked for statistical assumptions with diagnostic plots. Because the plots for both models suggested a skewed distribution of the residuals, we additionally assessed symmetry using the *skewness*() function from the *e1071* package v.1.7-16 ^37^ which yielded approximately symmetrical distribution (both, <0.2) of the residuals, and hence no violation of assumptions.

Marginal and conditional R^2^ were calculated as measures of goodness of fit for mixed models, in which marginal R^2^ (R^2^ _m_) reflects variance explained by fixed factors, and conditional R^2^(R^2^ _c_) the variance explained by the entire model. For the estimation of the main effects of the predictors, an analysis of variance (ANOVA) with Satterthwaite approximation for degrees of freedom was calculated on the models with the *anova*() function from base R. Post-hoc tests were performed with the *emmeans* package v.1.11-1 ^38^, with Holm correction.

In addition to the analyses described above, we re-run all the analyses for the sub-sample of participants left after removing eight participants (five AUT and three NT) who had less than 10 trials left after preprocessing in either of the two tasks (final sample N=66). The pattern of results remained unchanged in this sub-sample and hence we do not discuss it further in this article (the code and data accompanying this article and available in the OSF repository allow to reproduce both sets of analyses).

## 3. Results

### 3.1. Model 1: Group x Task

The distribution of the data across groups and tasks is shown in Figure 1A. First, we built a model with Group, Task, and their interaction as predictors (R^2^ _c_ = 0.67, R^2^ _m_ = 0.04). This model yielded a main effect of Task with steeper slopes in the active vs passive task, *F*(1, 6015.3) = 36.81, *p* < .001, BF10 > 1000 (very strong), a main effect of Group with steeper slopes in the NT than AUT group, *F*(1, 71.8) = 4.18, *p* = .045, BF10 = 1.6 (anecdotal), and a significant Group × Task interaction, *F*(1, 6015.3) = 27/2, *p* < .001, BF10 > 1000 (very strong). Post-hoc tests revealed that the interaction was driven by steeper slopes in active vs. passive task in AUT, *est*. = 0.06, *p*_*corr*_ < .001. No other pairwise comparisons were significant after corrections (see Supplementary Materials section 1). Figure 1B shows predictions of Model 1.

**Figure 1.**
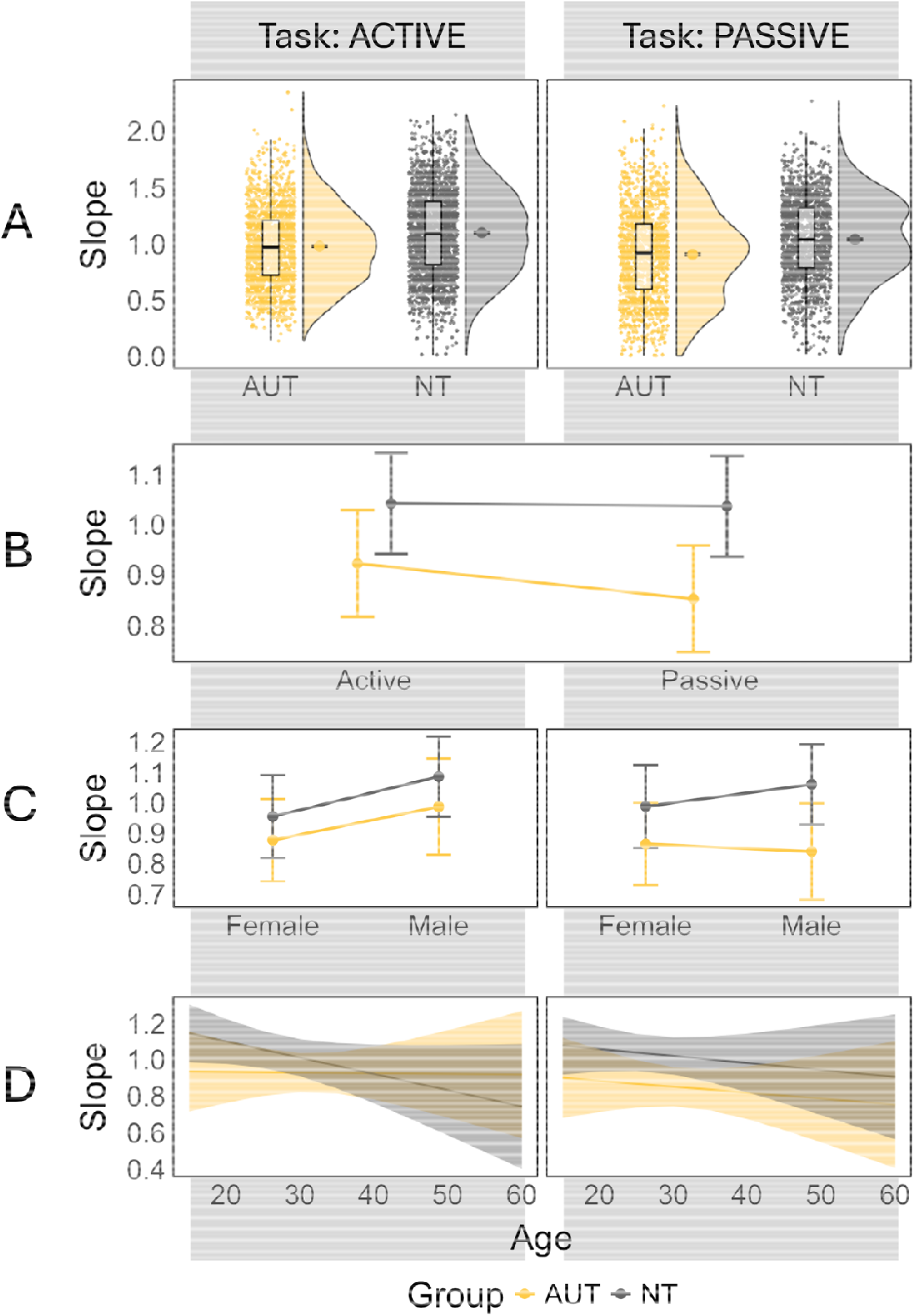
Group, task, age, and sex effects on aperiodic slopes. (A) Exponents of aperiodic slopes by group (autistic = AUT / neurotypical = NT) and task (active / passive). The point marks groups’ means with standard errors. (B) Model 1’s predictions of the group and task effects on the slopes. (C) Model 2’s predictions of the group, task, and sex effects. (D) Model 2’s predictions of the group, task, and age effects. In panels B, C, and D the ribbons mark 95% CI.

### 3.2. Model 2: Group x Task x Age x Sex

In the expanded model we added Age and Sex as predictors, together with their interactions with Group and Task (R^2^ _c_ = 0.69, R^2^ _m_ = 0.11). Adding Sex and Age significantly improved the model fit, *X*^*2*^(12) = 115.32, *p* < .001. As in the previous model, here we observed the main effect of Task, *F*(1, 6019.87) = 10.87, *p* < .001, BF10 > 1000 (very strong), and a Group x Task interaction, *F*(1, 6019.87) = 12.05, *p* < .001, BF10 > 1000 (very strong). Additionally, this model yielded two three-way interactions: Group x Task x Sex, *F*(1, 6019.87) = 4.15, *p* = .042, BF10 = 5.9 (moderate; see Figure 1C), and Group x Task x Age, *F*(1, 6021.00) = 33.89, *p* < .001, BF10 > 1000 (very strong; see Figure 1D). Post-hoc tests revealed that, as in Model 1, the Group x Task interaction was driven by significantly steeper slopes in active than passive task in the AUT group, *est*. = 0.08, *p*_*corr*_ < .001. The three-way interaction with Age was driven by a significantly stronger negative age effect in the NT group during the active vs. passive task, *est*. = -0.005, *p*_*corr*_ < .001, and a stronger positive age effect in AUT in the same task contrast, *est*. = 0.003, *p*_*corr*_ = .046. In the three-way interaction with Sex, no pairwise comparison of female vs. male survived corrections for multiple comparisons (see all statistics and post-hoc comparisons in Supplementary Materials, section 2).

### 3.3. Exploratory analysis – interindividual variability

To assess the robustness of our findings, we compared Model 2 (with a random intercept for participant) with Model 3, which additionally included a random slope for Task:

Model 3: Group x Task x Age x Sex + (1+Task|ID)

Including the random slope allows the model to account for interindividual variability in how participants respond to different tasks—that is, it captures the possibility that the effect of Task differs across individuals. This can improve the model’s flexibility and fit. Indeed, the model with the random slope provided a better overall fit, *X*^*2*^(2) = 1931.2, *p* < .001. However, under this model, there were no significant effects (see Supplementary Materials, section 3).

### 3.4. Exploratory analysis – trait covariates

As another exploratory analysis, we added AQ, LSAS, and IQ scores as covariates in Model 2. This did not improve the model, *X*^*2*^(3) = 5.63, *p* = .13. The main effect of interest – the Group x Task interaction – remained unchanged, and the only covariate which significantly predicted the aperiodic slopes was the LSAS score, so that the higher the score (i.e., the more social anxiety traits), the flatter the aperiodic slopes, *F*(1, 73.55) = 5.69, *p* = .02 (i.e., more excitation; for details, see Supplementary Material section 4).

## 4. Discussion

The present study investigated aperiodic slopes (the 1/f component) of the EEG signal in autistic and neurotypical adults during a passive viewing task compared to an active, goal-directed task. As expected, overall the slopes were steeper (indicating increased inhibition) in the active task compared to the passive one, consistent with previous findings ^14^. The key question here was whether this task effect would differ between groups (Group × Task interaction), although we had no specific directional hypotheses. The key finding to emerge from this study was that while neurotypical adults showed no task-related differences in aperiodic slopes, autistic adults showed significant increase (suggesting increased inhibition) during the active task relative to the passive one.

We hypothesised that a Group × Task interaction (rather than or in addition to a main effect of group), could indicate differences in how the autistic excitation/inhibition (E/I) systems dynamically respond to task demands compared to neurotypicals, rather than simply baseline differences. This interaction was indeed observed and was driven by significantly steeper slopes in the active compared to the passive task in the autistic group, while the neurotypical group showed no difference between tasks. Although not reaching statistical significance (anecdotal evidence in Model 1), the neurotypical group exhibited descriptively steeper slopes than the autistic group overall, consistent with the theory that autism is characterised by an E/I imbalance, typically reflecting elevated cortical excitation relative to inhibition ^2,6,39^.

Our findings therefore suggest that while autistic adults may exhibit a generally higher E/I ratio than their neurotypical peers, engaging in a more complex, goal-directed task may elicit a compensatory increase in inhibition—reflected in steeper aperiodic slopes—narrowing the gap between groups under active task conditions. This result is novel in the autism literature but aligns with broader evidence on the E/I system’s sensitivity to contextual demands, such as task engagement. Prior studies examining aperiodic slopes in autism have yielded mixed findings, ranging from steeper slopes in infants (but only among those with lower executive function; ^15^), to flatter slopes in children (but only in a below-average IQ sub-sample; ^16^), to no differences in adults ^19,40^. Here, we show that group differences may be exacerbated as a function of task context, in line with recent findings showing that the aperiodic slope can shift dynamically in response to externally induced changes in neural inhibition ^23^.

Controlling for age and sex did not affect the Group × Task interaction. However, we observed a three-way interaction among group, task, and age. This was driven by group differences in the active relative to the passive task: in the neurotypical group age was associated with significantly flatter slopes, whereas in the autistic group – with steeper slopes. This finding aligns well with previous work linking age to flatter slopes in the general population, though these effects have primarily been reported in resting-state conditions ^10,21^. Our results extend this literature by suggesting that age-related flattening may be more pronounced in active tasks, and it may be atypical in autistic individuals.

Collectively, the present findings highlight the importance of investigating the dynamic responsiveness of the autistic brain to task demands, particularly mechanisms that may increase inhibitory activity to compensate for elevated baseline E/I balance, rather than focusing solely on baseline group differences.

Given the well-established heterogeneity in autism ^41,42^, we explored interindividual variability in our data by adding random slopes for active/passive within participants to our main model. This adjustment improved model fit but rendered previously significant effects non-significant. Such a loss of significance likely reflects two factors: substantial individual differences in task responses, which require larger samples to detect consistent group effects, and the nesting of random slopes within participants, which absorbs variance that might otherwise be attributed to group. In other words, some variability previously modelled at the group level is better captured by individual differences, reducing power to detect group effects. This is particularly relevant for autism, where neural responses to cognitive demands may vary more widely than in typical development. The group-level Task × Group interaction seen in the simpler model may indicate a real trend, but the random-slope model reveals that it is not uniform across individuals. Modelling such variability is essential—not only for statistical precision, but also for representing the diversity of neural responses within the autistic population.

Several limitations should be noted. First, our design does not allow us to determine which specific aspect of the task drove the observed group differences in slopes. The contrast between passive and active conditions conflated multiple elements (presence vs absence of a behavioural goal, static vs dynamic stimuli, language-related vs non-linguistic input), and disentangling their relative contributions will require more targeted manipulations. Second, although our sample size was comparable to or larger than previous EEG studies in autism, the inclusion of random slopes suggested that substantial interindividual variability may have limited our ability to detect robust group-level effects. This points both to the need for larger datasets and to the value of analytic approaches that explicitly model within-group heterogeneity. Third, our study used only the visual-only condition of the active task to match the passive condition (see section 2.2 for the rationale behind this choice); other modalities (auditory or audiovisual) may reveal different dynamics. Finally, while we controlled for age and sex, other potentially relevant factors such as medication use, comorbidities, or circadian influences were not assessed, and may also shape aperiodic activity ^43^.

## 5. Conclusions

This study provides novel evidence that group differences in aperiodic slopes between autistic and neurotypical adults emerge primarily under task demands, rather than at rest. Autistic participants showed steeper slopes in the active compared to the passive task, consistent with a compensatory increase in inhibition when engaging in goal-directed behaviour. By contrast, neurotypical participants showed little change across conditions, though older age was associated with flatter slopes during active engagement. These findings suggest that aperiodic activity is not a static marker of group differences but a dynamic measure sensitive to context, task, and individual variability. More broadly, they highlight that task-related activity may reveal clinically relevant neural signatures not apparent in resting-state recordings, echoing similar observations in other conditions such as Parkinson’s disease or Alzheimer’s disease when controlling for vigilance state ^12^. Future studies with larger, more diverse samples and carefully matched task manipulations are needed to clarify the mechanisms and potential diagnostic utility of aperiodic EEG activity in autism.

## Data Availability

The data and analysis code will be made publicly available upon publication at OSF. For the review process, please use: https://osf.io/q64np/overview?view_only=5c059fbf30f44637bf762f2d871874c7.

## Acknowledgements

The authors thank the participants for their time and valuable insights and comments, and acknowledge the clinical autism centres in Catalonia, especially Autisme La Garriga, for their support in recruitment.

