## Supplementary Material for "Task-related Aperiodic EEG (1/f) Activity in Autism"

to:

### 1. Pairwise comparisons in Model 1

**Supp. Table 1** Pairwise comparisons for the effect of group and task interaction on aperiodic slopes (p.corr = with Holm correction).

|  | | | | | | |
| --- | --- | --- | --- | --- | --- | --- |
| **Contrast** | **estimate** | **SE** | **df** | **z.ratio** | **p.value** | **p.corr** |
| **Active** | | | | | | |
| AUT - NT | -0.118 | 0.074 | Inf | -1.601 | 0.109 | 0.219 |
| **Passive** | | | | | | |
| AUT - NT | -0.183 | 0.074 | Inf | -2.471 | 0.013 | 0.040 |
| **AUT** | | | | | | |
| Active - Passive | 0.070 | 0.009 | Inf | 7.559 | <0.001 | <0.001 |
| **NT** | | | | | | |
| Active - Passive | 0.005 | 0.008 | Inf | 0.642 | 0.521 | 0.521 |

### 2. Statistics and pairwise comparisons in Model 2

**Supp. Table 2** ANOVA results on Model 2 (Group x Task x Age x Sex)

|  | **Sum Sq** | **Mean Sq** | **NumDF** | **DenDF** | **F value** | **Pr(>F)** | **p.value** |
| --- | --- | --- | --- | --- | --- | --- | --- |
| **Group** | 0.069 | 0.069 | 1 | 73.819 | 1.328 | 0.253 | 0.253 |
| **Task** | 0.563 | 0.563 | 1 | 6019.869 | 10.869 | 0.001 | <.001 |
| **Age** | 0.073 | 0.073 | 1 | 73.768 | 1.409 | 0.239 | 0.239 |
| **Sex** | 0.169 | 0.169 | 1 | 73.819 | 3.266 | 0.075 | 0.075 |
| **Group:Task** | 0.624 | 0.624 | 1 | 6019.869 | 12.050 | 0.001 | <.001 |
| **Group:Age** | 0.021 | 0.021 | 1 | 73.768 | 0.402 | 0.528 | 0.528 |
| **Task:Age** | 0.135 | 0.135 | 1 | 6020.997 | 2.611 | 0.106 | 0.106 |
| **Group:Sex** | 0.015 | 0.015 | 1 | 73.819 | 0.294 | 0.589 | 0.589 |
| **Task:Sex** | 0.176 | 0.176 | 1 | 6019.869 | 3.405 | 0.065 | 0.065 |
| **Age:Sex** | 0.133 | 0.133 | 1 | 73.768 | 2.560 | 0.114 | 0.114 |
| **Group:Task:Age** | 1.756 | 1.756 | 1 | 6020.997 | 33.890 | 0.000 | <.001 |
| **Group:Task:Sex** | 0.215 | 0.215 | 1 | 6019.869 | 4.148 | 0.042 | 0.042 |
| **Group:Age:Sex** | 0.010 | 0.010 | 1 | 73.768 | 0.198 | 0.658 | 0.658 |
| **Task:Age:Sex** | 0.011 | 0.011 | 1 | 6020.997 | 0.207 | 0.649 | 0.649 |
| **Group:Task:Age:Sex** | 0.078 | 0.078 | 1 | 6020.997 | 1.504 | 0.220 | 0.22 |

Beside the main effect of Task, there are three significant interactions: Group x Task, Group x Task x Age, and Group x Task x Sex. Pairwise comparisons for all (with Holm corrections) are presented below.

Group x Task:

**Supp. Table 3** Pairwise comparisons for the effect of group and task interaction on slopes (with Holm correction)

| **contrast** | **estimate** | **SE** | **df** | **t.ratio** | **p.value** | **p.corr** |
| --- | --- | --- | --- | --- | --- | --- |
| **Active** | | | | | | |
| **AUT - NT** | -0.088 | 0.076 | 84.028 | -1.164 | 0.248 | 0.495 |
| **Passive** | | | | | | |
| **AUT - NT** | -0.172 | 0.076 | 84.827 | -2.261 | 0.026 | 0.079 |
| **AUT** | | | | | | |
| **Active - Passive** | 0.080 | 0.010 | 6026.467 | 8.311 | <0.001 | <0.001 |
| **NT** | | | | | | |
| **Active - Passive** | -0.004 | 0.008 | 6023.541 | -0.460 | 0.645 | 0.645 |

Group x Task x Age:

**Supp. Table 4** Pairwise comparisons of age slopes in all group and task combinations (with Holm correction)

| **contrast** | **estimate** | **SE** | **df** | **t.ratio** | **p.value** |
| --- | --- | --- | --- | --- | --- |
| **AUT Active - NT Active** | 0.009 | 0.008 | 84.406 | 1.096 | 1.000 |
| **AUT Active - AUT Passive** | 0.003 | 0.001 | 6033.091 | 2.608 | 0.046 |
| **AUT Active - NT Passive** | 0.004 | 0.008 | 84.749 | 0.458 | 1.000 |
| **NT Active - AUT Passive** | -0.006 | 0.008 | 84.518 | -0.735 | 1.000 |
| **NT Active - NT Passive** | -0.005 | 0.001 | 6022.252 | -6.273 | 0.000 |
| **AUT Passive - NT Passive** | 0.001 | 0.008 | 84.861 | 0.097 | 1.000 |

Group x Task x Sex:

**Supp. Table 5** Pairwise comparisons of sex differences in all group and task combinations (with Holm correction)

| **Contrast** | **Group** | **Task** | **estimate** | **SE** | **df** | **t.ratio** | **p.value** |
| --- | --- | --- | --- | --- | --- | --- | --- |
| **Female - Male** | AUT | Active | -0.110 | 0.112 | 84.349 | -0.979 | 0.331 |
|  | NT | Active | -0.131 | 0.102 | 83.644 | -1.281 | 0.204 |
|  | AUT | Passive | 0.025 | 0.112 | 84.942 | 0.218 | 0.828 |
|  | NT | Passive | -0.072 | 0.103 | 84.689 | -0.706 | 0.482 |

### 3. Statistics and pairwise comparisons in a model with random slopes for Task

**Supp. Table 6** ANOVA results on Model 3: Group x Task x Age x Sex + (1+Task|ID)

|  | **Sum Sq** | **Mean Sq** | **NumDF** | **DenDF** | **F value** | **Pr(>F)** |
| --- | --- | --- | --- | --- | --- | --- |
| **Group** | 0.023 | 0.023 | 1 | 65.114 | 0.630 | 0.43 |
| **Task** | 0.053 | 0.053 | 1 | 64.200 | 1.483 | 0.228 |
| **Age** | 0.056 | 0.056 | 1 | 65.000 | 1.569 | 0.215 |
| **Sex** | 0.097 | 0.097 | 1 | 65.114 | 2.702 | 0.105 |
| **Group:Task** | 0.000 | 0.000 | 1 | 64.200 | 0.001 | 0.98 |
| **Group:Age** | 0.002 | 0.002 | 1 | 65.000 | 0.066 | 0.799 |
| **Task:Age** | 0.023 | 0.023 | 1 | 64.132 | 0.640 | 0.427 |
| **Group:Sex** | 0.016 | 0.016 | 1 | 65.114 | 0.443 | 0.508 |
| **Task:Sex** | 0.000 | 0.000 | 1 | 64.200 | 0.000 | 0.991 |
| **Age:Sex** | 0.067 | 0.067 | 1 | 65.000 | 1.857 | 0.178 |
| **Group:Task:Age** | 0.011 | 0.011 | 1 | 64.132 | 0.293 | 0.59 |
| **Group:Task:Sex** | 0.000 | 0.000 | 1 | 64.200 | 0.002 | 0.967 |
| **Group:Age:Sex** | 0.013 | 0.013 | 1 | 65.000 | 0.351 | 0.556 |
| **Task:Age:Sex** | 0.004 | 0.004 | 1 | 64.132 | 0.118 | 0.732 |
| **Group:Task:Age:Sex** | 0.000 | 0.000 | 1 | 64.132 | 0.005 | 0.946 |

### 4. Exploratory analysis – trait covariates

**Supp. Table 7** ANOVA results on Model 2 with additional covariates: AQ, IQ, and LSAS.

|  | **Sum Sq** | **Mean Sq** | **NumDF** | **DenDF** | **F value** | **Pr(>F)** | **p value** |
| --- | --- | --- | --- | --- | --- | --- | --- |
| **Group** | 0.084 | 0.084 | 1 | 73.802 | 1.624 | 0.207 | 0.207 |
| **Task** | 0.562 | 0.562 | 1 | 6020.168 | 10.841 | 0.001 | <.001 |
| **Age** | 0.098 | 0.098 | 1 | 73.699 | 1.886 | 0.174 | 0.174 |
| **Sex** | 0.124 | 0.124 | 1 | 73.850 | 2.400 | 0.126 | 0.126 |
| **scale(AQ)** | 0.090 | 0.090 | 1 | 73.469 | 1.733 | 0.192 | 0.192 |
| **scale(IQ)** | 0.015 | 0.015 | 1 | 73.621 | 0.293 | 0.590 | 0.59 |
| **scale(LSAS)** | 0.295 | 0.295 | 1 | 73.545 | 5.687 | 0.020 | 0.02 |
| **Group:Task** | 0.623 | 0.623 | 1 | 6020.417 | 12.019 | 0.001 | <.001 |
| **Group:Age** | 0.054 | 0.054 | 1 | 73.757 | 1.044 | 0.310 | 0.31 |
| **Task:Age** | 0.134 | 0.134 | 1 | 6021.311 | 2.588 | 0.108 | 0.108 |
| **Group:Sex** | 0.024 | 0.024 | 1 | 73.839 | 0.457 | 0.501 | 0.501 |
| **Task:Sex** | 0.177 | 0.177 | 1 | 6020.148 | 3.418 | 0.065 | 0.065 |
| **Age:Sex** | 0.109 | 0.109 | 1 | 73.780 | 2.111 | 0.150 | 0.15 |
| **Group:Task:Age** | 1.753 | 1.753 | 1 | 6021.681 | 33.841 | 0.000 | <.001 |
| **Group:Task:Sex** | 0.214 | 0.214 | 1 | 6020.456 | 4.128 | 0.042 | 0.042 |
| **Group:Age:Sex** | 0.027 | 0.027 | 1 | 73.783 | 0.517 | 0.474 | 0.474 |
| **Task:Age:Sex** | 0.010 | 0.010 | 1 | 6021.116 | 0.199 | 0.655 | 0.655 |
| **Group:Task:Age:Sex** | 0.077 | 0.077 | 1 | 6021.729 | 1.495 | 0.222 | 0.222 |

### 5. Impact of segment length on aperiodic activity characterization

Given that passive trials were significantly shorter than visual-only trials for both groups (AUT: *t*(34) = -73.11, *p* < .001, NT: *t*(37) = -105.13, *p* < .001), we ran a control analysis in order confirm that the slope of aperiodic activity obtained in our experiment was not affected by segment length. For each subject, we simulated 500 datasets consisting of pink noise, with each dataset matching the segment length of the trials in the real experiment. For each subject and trial, we estimated the slope of simulated trials following the procedure described in the article. Next, for each subject the slopes were averaged across simulated conditions (simulated passive and simulated active) and compared with a paired two-tailed t-test with an alpha level of 0.05.

Mean slope values were 1.044±0.006 for the simulated passive trials and 1.036±0.006 for the simulated visual-only trials. The slope of simulated passive and simulated visual-only trials was significantly different in 9.2% of the simulations (in 0.2% of the simulations the slope of passive trials was flatter than visual-only trials, whereas in 9% of the simulations the slope of passive trials was steeper than visual-only trials). We used a Chi test in order to determine if the proportion of comparisons with a p-value below 0.05 differed significantly from the expected proportion, which was not the case, *X^2^*(1) = 0.0371, *p* = 0.8. Finally, we ran a Chi test on the number of significant p-values obtained for the simulation, comparing with a flat distribution. For the test, p-values were discretitzed in 20 bins. The distribution of p-values was not significantly different from the uniform distribution (*X^2^*(19) = 0.0771, *p* = 1).
